# Comparing immersive mindfulness and nature interventions for managing preoperative distress in breast cancer patients: A randomized controlled trial

**DOI:** 10.64898/2026.09.03.26362134

**Authors:** David Pinto, Rogélio Andrès-Luna, Adrian Razvan Sandru, Lydia Fettweis-Netto, Carlos Mavioso, Pedro Gouveia, João Correia-Anacleto, Luzia Travado, Patrícia Vaz, Alexandra Cruz, Zachary F Mainen, Maria João Cardoso, Scott Rennie

**Author notes:** These authors contributed equally to this work (David Pinto, Rogélio Andrès-Luna). Senior author (Scott Rennie).

## Abstract

Preoperative distress in breast cancer patients is highly prevalent and associated with adverse post-surgical outcomes. While both virtual reality (VR) distraction and mindfulness exercises have been increasingly utilized to manage this distress, the clinical impact of integrating active mindfulness meditation into the immersive experience remains under-investigated. Here, we present a randomized controlled trial evaluating the feasibility, acceptability and preliminary efficacy of brief, first-time immersive nature and mindfulness interventions for patients admitted to the holding area prior to the operating room for breast cancer surgery. Sixty participants were randomized to receive an 8-minute preoperative VR experience with mindfulness instruction (VR+Meditation), a standard VR nature experience without audio guidance (VR Only), or Care-as-Usual (CAU). The intervention was feasible and well tolerated with zero patient dropout and minimal indications of cybersickness or general discomfort. Outcomes included patient-reported anxiety and fear, physiological measures, and a computational semantic analysis of post-operative patient interviews. Compared with standard care, both VR interventions significantly reduced self-reported anxiety and heart rate. Although no significant differences were observed between the two active intervention groups, the VR+Meditation group demonstrated a significant and clinically meaningful reduction in surgery-specific fear relative to standard care. Analysis of brief patient interviews provided convergent evidence supporting these findings, showing that patients receiving VR+Meditation used language more strongly aligned with relief than those receiving standard care.

These findings suggest that immersive VR represents a feasible and well-tolerated first-time immersive intervention for reducing preoperative distress in patients undergoing breast cancer surgery. The addition of guided mindfulness may provide additional benefit by specifically addressing surgery-related fear, supporting the hypothesis that different digital interventions may target distinct dimensions of preoperative distress. Larger randomized studies are warranted to confirm these preliminary findings.

**Author summary:** Waiting for breast cancer surgery is an incredibly stressful experience, and high levels of anxiety before an operation can negatively impact a patient’s recovery and mental health. Because surgical wards are fast-paced, providing individualized psychological support in the moments just before an operation is challenging. In this study, we tested whether an immersive Virtual Reality (VR) experience may help reduce preoperative distress by transporting patients to a calming digital environment immediately before surgery. We compared standard VR distraction with the same immersive experience combined with guided mindfulness meditation. Both VR interventions reduced anxiety and heart rate compared with standard care. Only the group receiving VR with guided mindfulness showed a significant reduction in surgery-related fear and, when describing their experience after surgery, used language more strongly associated with relief.

Our findings suggest that immersive interventions provided to patients for the first time on the day of surgery can provide simple, well-tolerated psychological support during one of the most stressful moments in the patient journey. They also suggest that different digital approaches may influence different aspects of preoperative distress, with guided mindfulness offering additional support for surgery-related fear.

## Introduction

Breast cancer is the most common cancer in women worldwide with more than 2 million identified cases per year(1). Nearly all patients with early breast cancer undergo surgery as a component of treatment. Despite advances in surgical oncology and patient care, preoperative distress remains a pervasive issue, affecting a significant proportion of patients(2–4). In breast cancer patients, distress grows from the moment of diagnosis and frequently peaks during the pre-surgical phase.(5) Qualitative and quantitative data have reliably shown that both general anxiety and specific fear of surgery are major components of pre-surgical distress(2–4). This acute distress is important to address, not only as it is a profoundly negative experience during a vulnerable time in the patient journey, but also because high pre-operative distress is associated with adverse outcomes. This includes increased pain, nausea, delayed recovery, greater analgesic needs, and increased or new-onset depression(6–11).

The surgical context is a busy workplace designed to maintain sterility, safety and efficacy. While doctors and nurses work to provide a supportive patient experience this is often difficult due to these constraints and the fast paced nature of the environment. From the patient’s perspective this leaves them vulnerable, awaiting surgery in an unfamiliar, unpredictable and disempowering environment which can maintain or heighten the fear and anxiety associated with treatment(12–15). An intervention to support patients as they face pre-surgical distress in the perioperative environment must improve their experience of it without interfering with its medical requirements. Ideally it would also be possible to administer the intervention without prior patient experience, with minimal disruption to medical workflows or additional nurse training.

The central feature of VR is that it can transport users to new contexts creating a vivid and convincing sensation of being immersed in a simulated environment(16). This offers the possibility of reimagining the experience of the pre-operative space for patients without changing it avoiding the logistical, procedural and financial costs associated with the extensive redesign of clinical spaces or procedures. As a result VR’s has begun to emerge as a powerful non-pharmacological tool to support patient experience and reduce preoperative anxiety(17).

VR has generally been used to address preoperative anxiety in one of two ways, either to inform patients about what to expect on the day of surgery through immersive walk-throughs or to provide distraction by immersing them in calming environments that can override and reduce the anxiogenic reality of the medical context during the perioperative period. A recent meta-review examining the use of VR in patients undergoing elective surgery with anaesthesia has demonstrated that acute waiting room distraction-based approaches are more effective than using VR to inform and prepare patients prior to surgery in reducing preoperative distress in both pediatric and adult patients(18). Nonetheless the majority of evidence for VR distractions efficacy, while positive, is derived from pediatric patients(19,20). In adults meta analyses focusing on perioperative anxiety have shown mixed results with some showing moderate to large effect sizes of VR distraction in reducing pre-surgical distress and procedural pain(19,21–23), while others show low(17) to non-significant effects(24) when effect sizes are pooled.

Current research on VR distraction targets the efficacy of the technology itself, with little direct comparison of experiential content presented to participants. Most studies have evaluated whether VR works, but very few have compared how different therapeutic content delivered within the immersive environment influences clinical outcomes. This oversight is striking considering that the use of VR in the direct pre-surgical window to address anxiety depends upon predictably altering patient experience.. This study is the first to test whether established anxiolytic instruction enhance the beneficial effects of VR distraction.

Mindfulness meditation based therapies for anxiety are both effective and diverse. They range from structured multi-session treatments such as Mindfulness Based Stress Reduction (MBSR) and Mindfulness Based Cognitive Therapy (MBCT) to short first time acute interventions provided through audio recordings or an in-person guide(25). The common core across all these approaches is the self-regulation of attention maintained on a specific, immediate target, coupled with a curious and accepting orientation toward the experience of that focus(26).

Mindfulness-based interventions (MBI’s) have demonstrated the capacity to reduce pre-operative distress reported experience and physiological markers, lowering both cortisol and heart rate(27–30).

However, anxiety is known to impair attentional control. While brief first time interventions are practical and can be effective in the pre-surgical context(31), this tends to depend on patients predisposition towards or experience with mindfulness(29). Nonetheless the less first time interventions tend to be less effective than longer structured training programmes like MBSR especially in high anxiety groups or high-risk patients such as oncology patients (25,27,30,32). This is likely because the anxiogenic aspects of the surgical context makes it difficult for untrained participants without in-person guidance to maintain attentional focus. VR may address this by creating a curated immersive experience, alongside audio guidance, that limits exposure to anxiogenic aspects of the waiting room and that supports attentional control (33). Early research already supports this, demonstrating that a repeated, multi-session perioperative VR mindfulness protocol is safe, feasible, and yields high patient satisfaction, however it has only been tested as a repeated intervention and compared to care as usual (34).

Combining VR distraction with active mindfulness guidance may improve the effectiveness of brief first time mindfulness interventions without the expense of in-person professional guidance or more extensive long-term training. This paper reports on a pilot randomized controlled trial designed primarily to establish the feasibility and acceptability of deploying a brief, combined VR-mindfulness intervention for pre-operative breast cancer patients within a clinical setting. Secondarily, we aimed to evaluate the preliminary efficacy of these brief, first-time immersive interventions for pre-operative breast cancer patients. We hypothesized that combining immersive VR with guided mindfulness would provide greater reductions in preoperative distress than VR distraction alone or standard care. Specifically, we evaluated the effects of these interventions on patient-reported anxiety and surgery-specific fear, physiological responses, and postoperative recollections of the preoperative experience.

## Methods

### Recruitment

Participants were approached during pre-operative consultations, where they were provided with detailed information regarding the study’s objectives and procedures. A total of sixty female patients scheduled for elective breast cancer surgery at the Champalimaud Foundation were recruited for this study (fig 1.a) from 2/1/2024 until 31/3/2025. Inclusion criteria required participants to be aged 18 years or older, capable of providing written informed consent, and indicated for surgical treatment requiring general anaesthesia.

**Figure 1:**
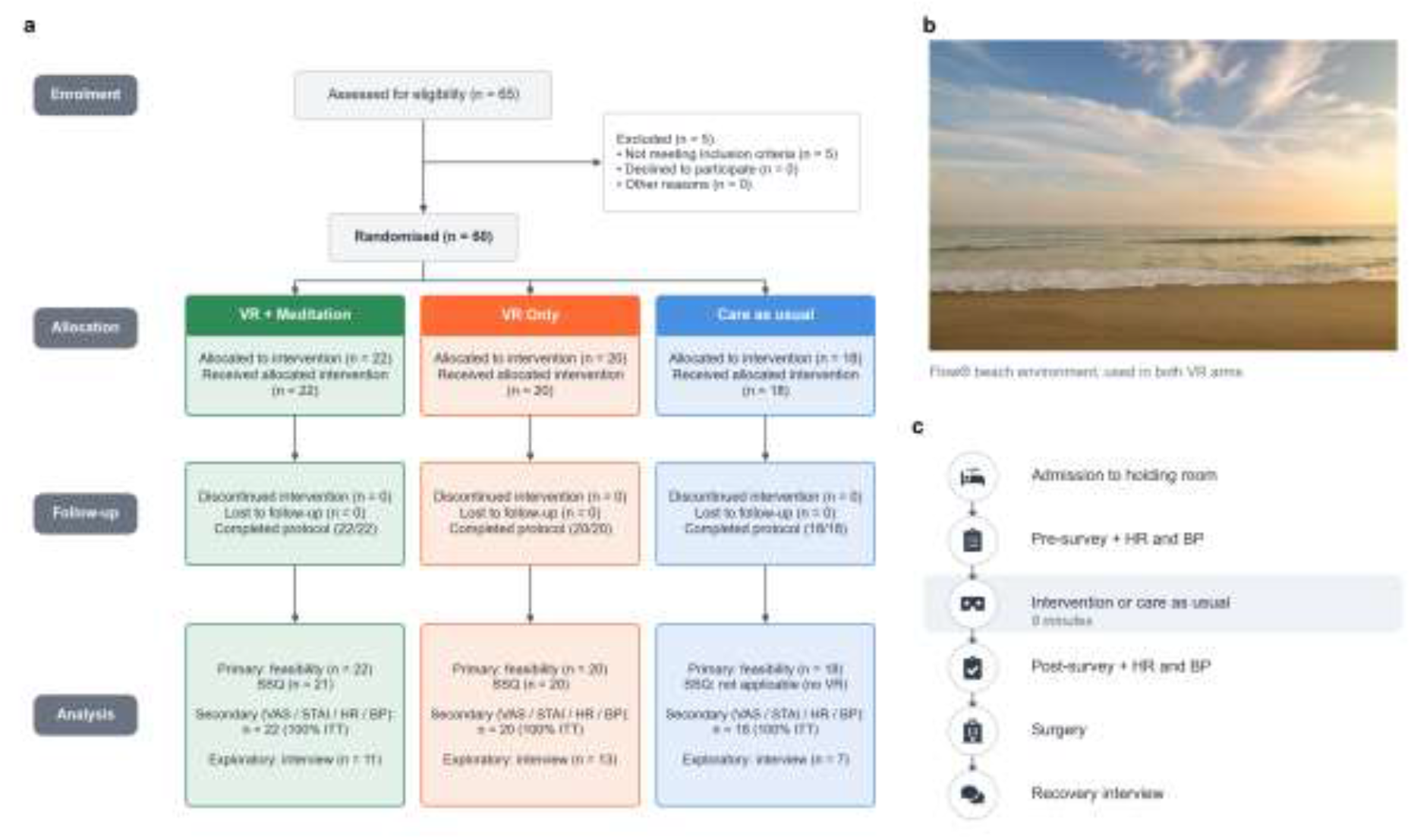
Study Design, Virtual Environment, and Procedural Timeline. **(a) Participant Flow Diagram:** Sixty patients scheduled for elective breast cancer surgery were recruited and randomized into three study arms: VR+Meditation (n=22), Care As Usual (n=18), and VR Only (n=20). The diagram outlines the number of participants per group with recorded physiological data (Heart Rate and Blood Pressure) and those who participated in the exploratory post-operative recovery interviews. All sixty randomized participants were included in the final analysis. **(b) Immersive Virtual Environment:** A representative screenshot of the naturalistic beach environment utilized in the Flow® application. This visual stimulus was presented to participants in both the VR+Meditation (with audio guidance) and VR Only (without audio guidance) intervention arms. **(c) Perioperative Procedure Timeline:** Upon admission to the preoperative holding room, patients completed baseline surveys alongside initial heart rate and blood pressure measurements. Participants then underwent their assigned 8-minute intervention (VR+Meditation, VR Only, or CAU). Immediately following the 8-minute period, post-intervention surveys and physiological metrics were collected before patients were transferred to the operating theatre. Optional semi-structured interviews were conducted during post-operative recovery.

Exclusion criteria comprised severe visual or auditory impairments, cognitive deficits that would preclude engagement with the VR hardware, and a history of motion sickness or known contraindications to VR exposure.

#### VR Hardware and Software description

Flow^®^ is a commercially available immersive meditation application, that integrates real nature visuals (fig.1.b). We developed a specific study version from the product commercially available. Flow^®^ installed this study version in a VR headset Pico Neo 3 Head-Mounted Display that was used for the intervention arms. Following consultation with a patient advocate, a beach environment was selected for both intervention arms. The English meditation script was translated into Portuguese for the study.

#### Study Design

This investigator-initiated pilot randomized controlled trial was designed by the authors in collaboration with a Portuguese patient advocate. We conducted a single-cohort, prospective randomized study consisting of three arms (allocation 1.1.1). To ensure baseline equivalence, participants were randomly assigned to one of three study arms using a computerized random number generator. The conditions were defined as follows:

1. **VR + Meditation:** Participants experienced an 8-minute guided meditation using the Flow^®^ VR meditation application. This included an immersive virtual beach environment designed to evoke relaxation, combined with a guided breath meditation translated into Portuguese.
2. **VR Only (Active Control):** Participants experienced the identical 8-minute immersive beach environment without the guided meditation auditory component, serving as a standard VR distraction treatment.
3. **Care-as-Usual (CAU):** Participants received standard pre-operative care in the same environment for an equivalent 8-minute period without any VR exposure, music or human interaction.

### Trial Registration and Outcome Measures

This trial was registered retrospectively on ClinicalTrials.gov (Identifier:NCT07785245). Prospective registration was not completed prior to patient enrollment due to the initial exploratory focus on feasibility. To align with the trial’s registration and pilot design, outcomes were evaluated hierarchically:

#### Primary Outcome (Feasibility & Acceptability)

The primary objective was to establish the viability of the VR interventions in the immediate pre-operative setting. This was evaluated via participant drop-off/discontinuation rates (with a predefined threshold of <10% for success) and overall patient tolerability and acceptance, measured using an adapted Simulator Sickness Questionnaire (SSQ) and a recommendation scale administered immediately post-intervention.

#### Secondary Outcomes (Preliminary Clinical Efficacy)

Secondary objectives assessed the psychological and physiological efficacy of the interventions. Psychological measures included changes in state anxiety (assessed via the short Brazilian State Trait Anxiety Inventory [STAI-S] and a 0-100 Visual Analogue Scale [VAS]), surgery-specific fear (VAS Afraid), and mood (VAS Calm and VAS Optimistic). Physiological arousal was measured via continuous monitoring of resting heart rate and blood pressure. All secondary measures were recorded at baseline and immediately following the 8-minute intervention.

#### Exploratory Outcome (Qualitative Sentiment)

To capture a more nuanced understanding of the patient experience, exploratory post-operative semi-structured interviews were conducted. Transcripts were subjected to natural language processing and semantic projection analysis to quantify subjective alignment along a distress-to-relief axis.

#### Perioperative Procedure and Data Collection

On the day of surgery, within the pre-operative room inside operating room facilities, patients underwent standard clinical preparation by a nurse, including peripheral vascular access and standard monitoring of blood pressure, pulse oximetry and heart rate. Directly prior to the intervention, participants completed the pre-session survey (fig.1.c) which included Visual Analogue Scales (VAS) assessing Anxiety, Fear, Optimism, and Calmness, the short Brazilian State Trait Anxiety index(35), four VAS items taken from the Simulator Sickness Questionnaire (SSQ)(36). These were administered in Portuguese via Qualtrics on an Android tablet.

Following the initial assessment, participants in the two VR conditions were fitted with the VR headset for the 8-minute intervention, while the CAU group rested for the same duration. Immediately after the allotted time had elapsed, post-intervention physiological metrics (heart rate and blood pressure) were manually recorded from the continuous monitors.

Participants then completed the post-intervention survey on an android tablet, which for the VR groups included an additional adapted SSQ to measure patient tolerability and incidence of cybersickness. Following the completion of this final assessment, patients were transferred directly to the operating theatre. Following recovery, a post-operative semi-structured interview was carried out by attending surgeons about their experience.

#### Semantic analysis

Interviews were conducted according to patient availability and constrained medical scheduling, which led to a total of 32 interviews. One Interview was excluded from the analysis due to lack of state relevant vocabulary. The interviews were audio recorded, then transcribed, diarized and translated using WhisperX (3.4.2)(37). All transcripts and translations were human verified by a Portuguese native speaker. Psychological and experiential state descriptors were extracted by a native Portuguese speaker and cross-checked by a second researcher. We quantified language along a predefined semantic axis spanning distress to relaxation and tested whether participants differed in position within the semantic subspace defined by the semantic axis based on experimental condition.

Following the semantic projection framework described by(38), semantic representations were derived using a Sentence-Transformers pipeline built from *Qwen/Qwen3-Embedding-0.6B*. This model was selected because of its strong performance on the MTEB STS benchmark and its support for instructed inputs, which were used here to better capture the valence and intensity of each word or short phrase. As robustness checks, the data were also embedded using the original *GloVe 300* embedding space used by Grand *et al*. (2022) (38), as well as *jina-embeddings-v5-text-small*, a high-performing model on the MTEB STS benchmark, like Qwen, but based on a different architecture. A distress-relaxed semantic axis was constructed from anchor terms indexing distress (“distressed”, “danger”, “apprehensive”, “anguish”, “torment”, “agitated”) and relaxation (“relieved”, “comforted”, “security”, “soothed”, “reassured”, “at ease”). The anchor terms were defined based on―distress‖ and ―relaxed‖ synonyms that were not used by the participants, to avoid circularity between the semantic axis and the participant-generated vocabulary being projected onto it. Consistent with the semantic projection procedure, the axis was defined as the mean of all pairwise difference vectors between distress and relaxation anchor embeddings. Each target word was then assigned a scalar projection score on this axis, computed as (u · v) / ||u||, where u denotes the distress-relaxation axis vector and v denotes the embedding of the target word on the axis. Lower scores reflected greater semantic alignment with distress, whereas higher scores reflected greater semantic alignment with relaxation.

#### Statistical methods

All analyses were conducted using Python 3.12.12 with the following packages: SciPy (stats module) for basic statistical tests, statsmodels for ANOVA/ANCOVA/MANOVA models, NumPy for numerical computations, and pandas for data manipulation. Visualization was performed using matplotlib and seaborn. All analysis code is available upon request to support reproducibility.

#### Tolerability and Cybersickness

Cybersickness symptoms (Nausea, Dizziness, Headache, Blurred Vision) were assessed using items adapted from the Simulator Sickness Questionnaire on a 0–100 scale, administered to VR groups only. Descriptive statistics (mean, SD) were computed per symptom per group. Between-group differences (VR+Meditation vs VR Only) were evaluated using independent-samples t-tests and Mann-Whitney U tests (two-tailed). An overall single-item discomfort rating (“How uncomfortable was the experience?”, 0–100) was also reported, with the proportion of participants scoring above 40/100 noted as an indicator of clinically meaningful discomfort.

#### Visual Analogue change scores

Multivariate analysis employed a one-way multivariate analysis of covariance (MANCOVA) to assess the overall treatment effect across all four VAS emotional measures (Anxious, Optimistic, Afraid, Calm) while controlling for their respective baseline values. Specifically, pre-intervention VAS scores for each measure (VAS Anxious Pre, VAS Optimistic Pre, VAS Afraid Pre, and VAS Calm Pre) were included as covariates in the model. Wilks’ lambda was used as the multivariate test statistic, with partial eta-squared (η²) reported as the measure of multivariate effect size.

Follow-up one-way ANOVAs were conducted on change scores (post-intervention minus pre-intervention) for each VAS outcome to identify specific measures driving the multivariate effect. Effect sizes were quantified using eta-squared (η²), with values of 0.01, 0.06, and 0.14 representing small, medium, and large effects, respectively.

#### Post-hoc Comparisons

Pairwise comparisons between treatment groups were conducted using independent-samples t-tests with Bonferroni correction (p-values multiplied by the number of comparisons) to control for Type I error inflation. Effect sizes for pairwise comparisons were calculated using Cohen’s d, with pooled standard deviation. Cohen’s d values of 0.2, 0.5, and 0.8 were interpreted as small, medium, and large effects, respectively. Bootstrap confidence intervals (95%) were computed for Cohen’s d estimates to account for potential departures from normality.

#### Heart Rate Analysis

Heart rate change scores (post-pre) were analyzed using one-way ANOVA with follow-up pairwise comparisons to evaluate physiological treatment effects. Given the non-normal distribution of HR change scores in the VR Only group (driven by a single extreme value), non-parametric tests (Mann-Whitney U, Wilcoxon signed-rank) were reported alongside parametric results.

#### Predictive Analyses

Multiple regression analyses were conducted to examine whether baseline characteristics (e.g., trait anxiety measured by STAI-T) predicted treatment response, with VAS change scores as outcomes and treatment condition included as a categorical predictor. As an exploratory analysis, change scores in VR- and meditation-naïve patients (prior VR or meditation rating below 10/100) were compared with care as usual using Welch t-tests.

Clinical significance was assessed using the established minimal clinically important difference (MCID) of 10 points for VAS scales (0-100 scale). Participants achieving a change score ≥10 points in the therapeutic direction (reduction for negative emotions, increase for positive emotions) were classified as responders. Response rates were compared across treatment groups using Fisher’s exact test for pairwise comparisons. Number Needed to Treat (NNT) was calculated as the reciprocal of the absolute risk difference between intervention and control conditions. NNT values indicate the number of patients who need to receive the intervention to produce one additional responder compared to control. As these responder analyses were exploratory and secondary to the primary MANCOVA analysis, no alpha-level corrections (e.g., Bonferroni) were applied to the Fisher’s exact test p-values.

### Statistical Assumptions and Robustness

#### Normality Testing

Shapiro-Wilk tests were conducted to assess normality of change score distributions. Where departures from normality were detected, bootstrap confidence intervals were computed to ensure robust inference.

#### Outlier Analysis

Outliers were identified using the interquartile range (IQR) method (values beyond 1.5 × IQR from the first or third quartile). The prevalence of outliers was reported for each outcome and treatment group. Primary analyses included all participants (intent-to-treat principle), with sensitivity analyses conducted excluding outliers when appropriate.

#### Missing Data

Complete case analysis was employed, with participants included in analyses only for outcomes with complete pre- and post-intervention data. The proportion of missing data was reported for each outcome measure, and patterns of missingness were examined to assess potential bias.

#### Semantic analyses

Conducted using Python 3.12.12 and R. Semantic embedding and preprocessing were implemented in Python using sentence-transformers (Version 5.2.2), transformers (Version 5.0.0), torch (Version 2.9.0), scikit-learn (Version 1.6.1), numpy (Version 2.0.2), pandas (Version 2.2.2), and matplotlib (Version 3.10.0). Inferential analyses were conducted in R using the lme4, lmerTest, clubSandwich, performance, and ggplot2 packages.

### Ethics and Data Management

The study protocol was reviewed and approved by the institutional ethics board at the Champalimaud Foundation. All participants provided explicit informed-consent prior to being enrolled in the study. All collected data were rigorously anonymised, securely stored on password-protected digital devices, and handled in strict compliance with institutional and international data protection standards

### Availability of Data and Materials

The anonymized dataset analyzed during the current study and the corresponding analytical code (including the NLP workflow and semantic projection scripts) are available in two public repositories. https://github.com/SMckenzieRennie/preop-vr-analysis, https://github.com/arsandru/preop-vr-NLP

## RESULTS

Sixty adult female patients scheduled for elective surgery were recruited and randomized into one of three groups. VR+Meditation (n=22), VR Only (n=20), or Care as Usual control (n=18). Randomization was stratified by the Champalimaud clinical Trials team. Inclusion criteria comprised sufficient vision and hearing to engage with the VR experience and the ability to provide informed consent. Exclusion criteria included a history of motion sickness or contraindications to VR exposure. Participants and assessors were necessarily unblinded to treatment conditions.

Participants’ mean age was 55.9 years (SD = 11.6, range 26-85), with no significant age differences between treatment groups (*F*(2,57) = 0.02, p = .980). Surgical procedures included breast-conserving surgery (25), breast-conserving surgery with symmetrization (24), mastectomy with immediate reconstruction with or without symmetrization (8), other reconstructive procedures (2), and prophylactic surgery (1) (see Table 1). The majority of participants (58.3%) were scheduled for surgery as first cancer treatment, while 40.0% were scheduled for neoadjuvant chemotherapy followed by surgery. Approximately 41.7% of participants had prior breast surgery or cancer treatment history.

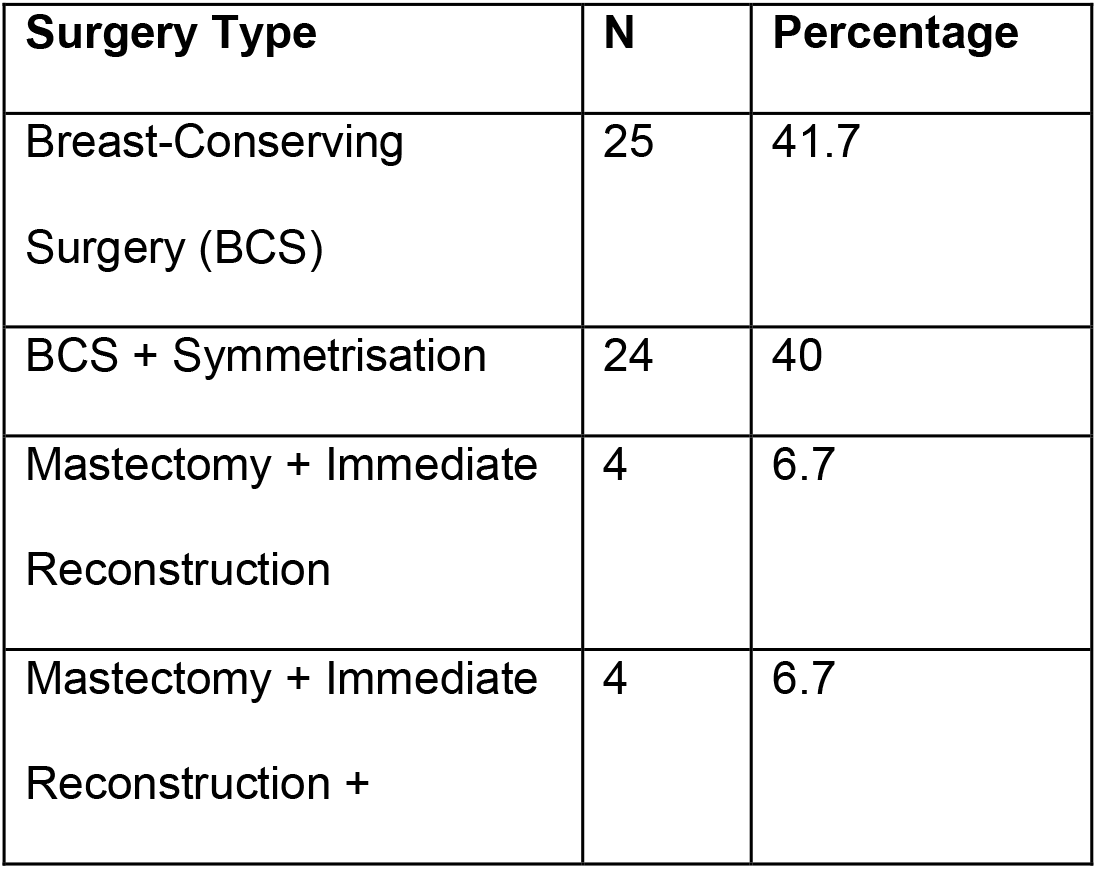
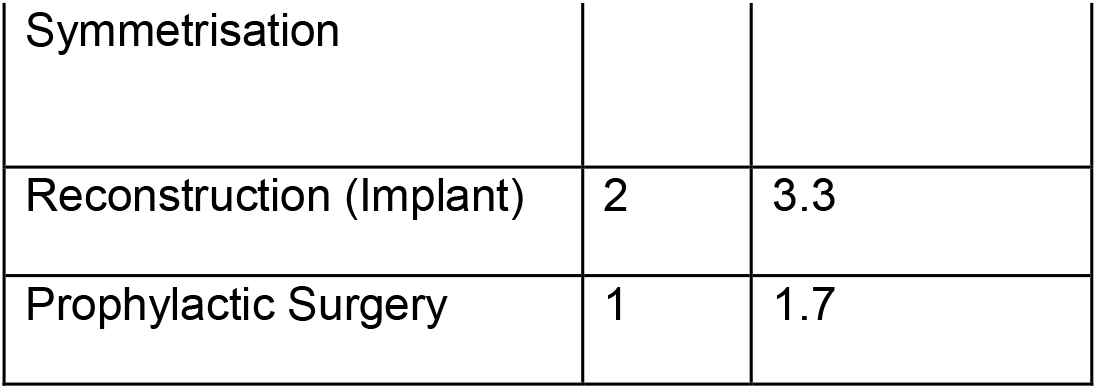

### Measures

At baseline, participants reported moderate anxiety levels across measures: STAI-S (M = 15.4, SD = 1.7, range 10-19), VAS Anxious (M = 50.9, SD = 27.9, range 0-100), and VAS Afraid (M = 43.1, SD = 31.2, range 3-100).

| Baseline Measure | VR+Meditation M (SD) | VR Only M (SD) | Control M (SD) | Overall M (SD) | Range |
| --- | --- | --- | --- | --- | --- |
| STAI-S Pre | 15.5 (1.3) | 15.6 (1.5) | 15.1 (2.3) | 15.4 (1.7) | 10-19 |
| VAS Anxious Pre | 45.9 (30.3) | 52.7 (27.1) | 55.0 (26.5) | 50.9 (27.9) | 0-100 |
| VAS Optimistic Pre | 84.0 (15.5) | 82.3 (18.3) | 83.8 (17.2) | 83.4 (16.7) | 32-100 |
| VAS Afraid Pre | 46.8 (31.6) | 35.0 (28.7) | 47.7 (33.2) | 43.1 (31.2) | 3-100 |
| VAS Calm Pre | 43.5 (26.8) | 53.6 (24.9) | 48.1 (23.7) | 48.2 (25.2) | 3-99 |

The primary outcomes measures were feasibility and acceptability. Both VR interventions were well tolerated, and no participants discontinued treatment due to discomfort. Cybersickness reports were minimal. The VR+Meditation composite mean was 2.15 (SD = 1.83; complete n = 21; range 0–5.75), and the VR Only composite mean was 3.54 (SD = 4.77; complete n = 20; range 0–20.75). All evaluable participants scored well below the pre-defined clinical acceptability threshold of < 25: 21/21 (100%) in VR+Meditation and 20/20 (100%) in VR Only. The between-group difference for the composite score was not significant via Mann–Whitney U test (U = 185.5, p = .531). One VR+Meditation participant had incomplete four-item symptom data and was excluded from this test. Individual symptom-level comparisons were also non-significant (all Welch p ≥ .090 and all Mann–Whitney p ≥ .070) (table.3). Additionally, participants rated both experiences as highly relaxing and engaging (means > 77 on the 0–100 scale). The single-item ―Uncomfortable‖ rating was also lower for VR+Meditation than VR Only: 5.6 ± 7.0 versus 16.6 ± 23.6 (table.4).

**Table.3.**

| Symptom (0-100) | VR+Meditation M (SD) | VR Only M (SD) |
| --- | --- | --- |
| Nausea | 2.0 (2.1) | 3.2 (2.5) |
| Dizziness | 2.0 (2.1) | 3.1 (3.6) |
| Headache | 2.0 (1.7) | 3.8 (6.7) |
| Blurred Vision | 2.3 (2.6) | 4.0 (7.4) |
| Composite score | 2.15 (1.83) | 3.54 (4.77) |

**Table.4.**
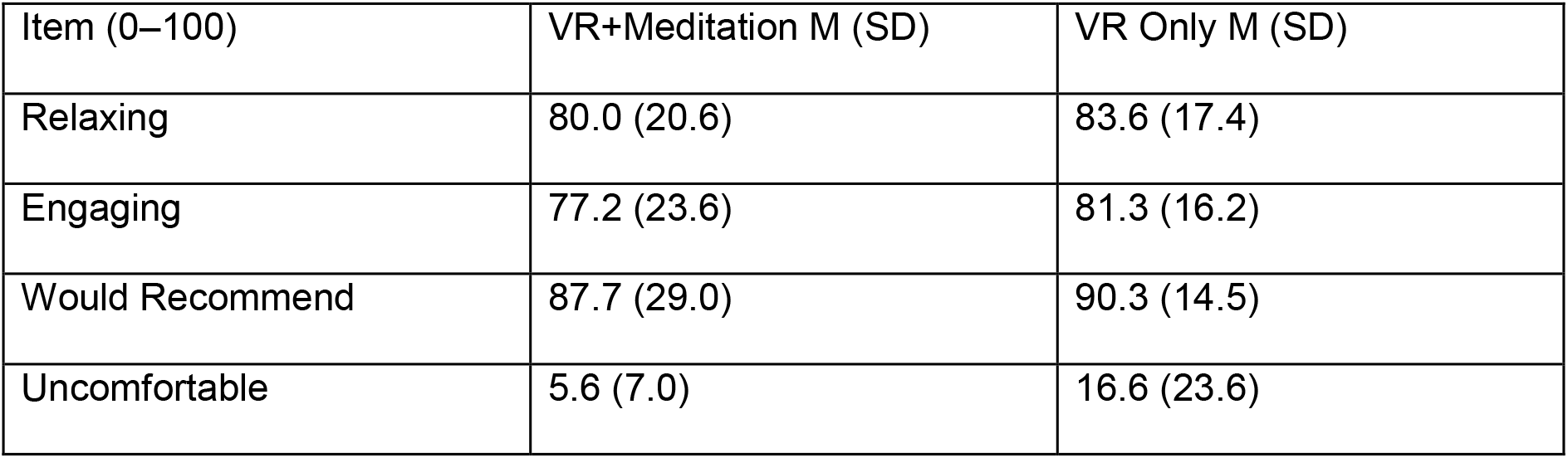

| Item (0–100) | VR+Meditation M (SD) | VR Only M (SD) |
| --- | --- | --- |
| Relaxing | 80.0 (20.6) | 83.6 (17.4) |
| Engaging | 77.2 (23.6) | 81.3 (16.2) |
| Would Recommend | 87.7 (29.0) | 90.3 (14.5) |
| Uncomfortable | 5.6 (7.0) | 16.6 (23.6) |

### Secondary Outcomes – Psychological Efficacy

A one-way MANCOVA controlling for baseline VAS scores revealed a significant overall treatment effect across the combined post-intervention VAS scores (Wilks’ λ = 0.655, F(8, 100) = 2.95, p = 0.005, η² = 0.345), demonstrating an effect of treatment upon VAS responses. Follow-up univariate ANOVAs showed significant group effects for Anxious [F(2, 57) = 5.04, p = .010] and Afraid [*F*(2, 57) = 5.48, p = .007] ratings (fig.2.a-d). No significant differences between treatment groups were found for Calm or Optimistic. Pairwise comparisons with Bonferroni correction revealed that VR+Meditation (p = 0.016) and VR Only (p = 0.006) significantly outperformed Care as Usual, with VR Only showing the largest effect size (Cohen’s d = −1.070). For Fear, only VR+Meditation showed a significant reduction relative to Care as Usual (p = 0.007, Cohen’s d = −0.991), while VR Only showed a moderate but non-significant improvement (p = 0.185, Cohen’s d = −0.621). Direct comparisons between VR+Meditation and VR Only did not reach statistical significance for either measure (both p > 0.30),(table 5). These findings suggest that pre-surgical distress comprises separable psychological components general anxiety and surgery specific fear. While standard immersion in nature is sufficient to alleviate generalized anxiety, the integration of guided mindfulness appears to uniquely act upon surgery-specific fear

**Figure 2:**
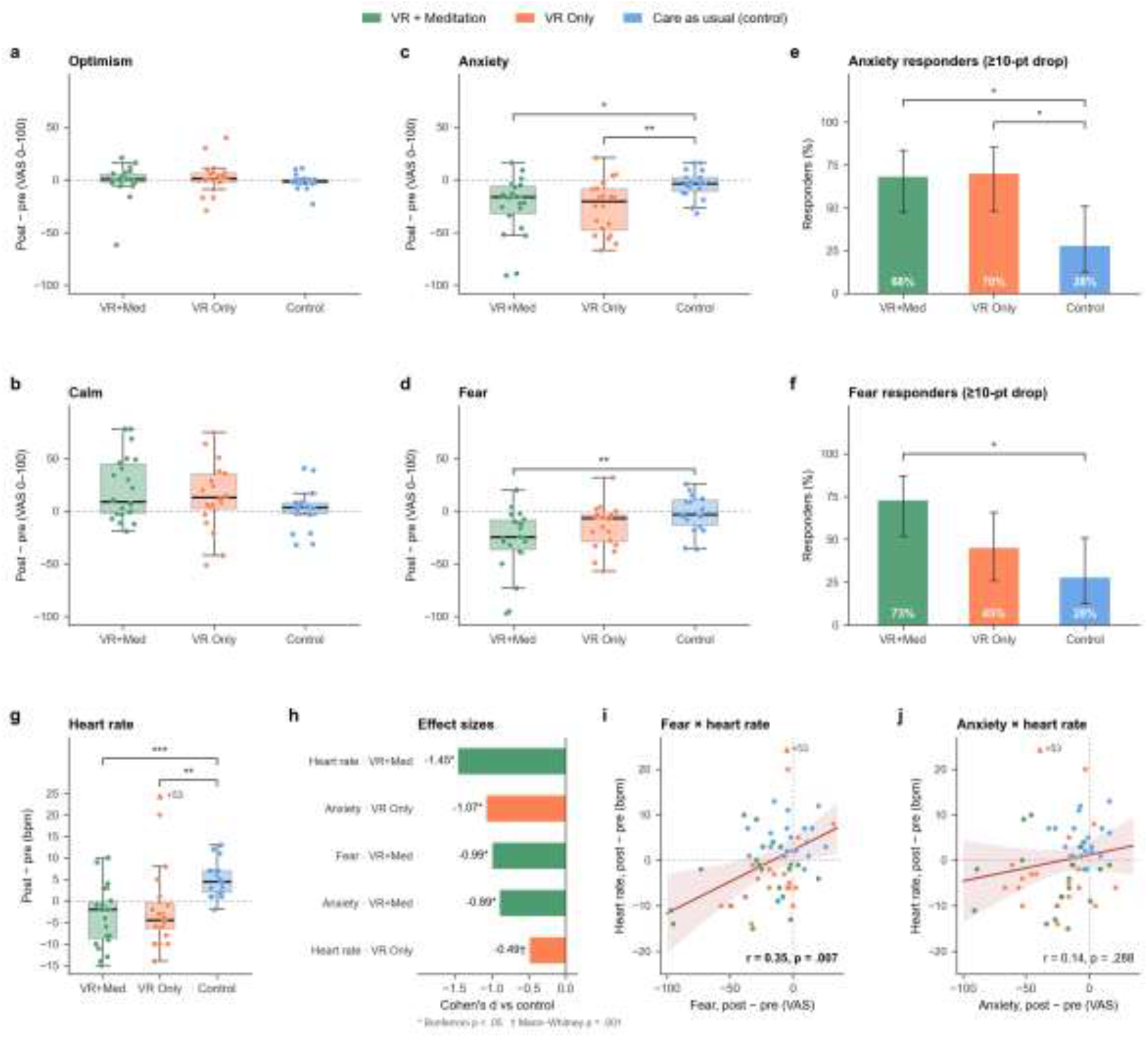
Clinical and Physiological Impact of Virtual Reality and Guided Meditation on Preoperative Distress. **(a-d) VAS change:** No significant difference between groups was found for **(a)** Optimistic or **(b)** Calm; however, **(c)** both VR+Meditation (p=0.016) and VR Only (p=0.006) significantly reduced anxiety vs. control, and **(d)** VR+Meditation significantly reduced fear vs. control (p=0.007).**(e-f)** Responders by VAS scores: High clinical response rates (≥10-point reduction) for VR+Meditation (68.2%) and VR Only (70.0%) vs. control (27.8%) were found for **(e)** anxiety, and for **(f)** fear, VR+Meditation (72.7%) outperformed both VR Only (45.0%) and control (27.8%). **(g)** Heart Rate Change: Both VR+Meditation and VR Only interventions significantly reduced heart rate compared to the control group. **(h)** Effect Sizes: Cohen’s d confirms large treatment effects of the VR interventions for anxiety, fear, and heart rate reductions vs. control. **(i-j)** Correlations between HR and VAS change scores: Fear **(i),** but not anxiety **(j)**, change scores correlate with pre-post changes in heart rate across the whole cohort.

**Table 5.**

| Measure / Timepoint | VR+Meditation (n=22) | VR Only (n=20) | Control (n=18) | Group Effect & Post-hoc (vs. Control) |
| --- | --- | --- | --- | --- |
| **VAS Anxious** |  |  |  | **F(2,57)=5.04, p=0.010** |
| Pre (M ± SD) | 45.9 ± 30.3 | 52.7 ± 27.1 | 55.0 ± 26.5 | |
| Post (M ± SD) | 21.6 ± 18.5 | 26.7 ± 26.8 | 50.6 ± 27.8 | |
| Cohen's d | d=-0.89 | d=-1.07 | — | VR+Med: p=0.016,<br>VR Only: p=0.006 |
| **VAS Afraid** |  |  |  | **F(2,57)=5.48,** **p=0.007** |
| Pre (M ± SD) | 46.8 ± 31.6 | 35.0 ± 28.7 | 47.7 ± 33.2 |  |
| Post (M ± SD) | 19.6 ± 19.3 | 20.8 ± 18.6 | 45.4 ± 26.7 |  |
| Cohen's d | d=-0.99 | d=-0.62 | — | VR+Med: p=0.007,<br>VR Only: p=0.185 |
| **VAS Calm** |  |  |  | **F(2,57)=2.39,**<br>**p=0.101** |
| Pre (M ± SD) | 43.5 ± 26.8 | 53.6 ± 24.9 | 48.1 ± 23.7 |  |
| Post (M ± SD) | 64.8 ± 31.2 | 68.3 ± 28.8 | 50.1 ± 24.3 |  |
| **VAS Optimistic** |  |  |  | **F(2,57)=0.42,**<br>**p=0.658** |
| Pre (M ± SD) | 84.0 ± 15.5 | 82.3 ± 18.3 | 83.8 ± 17.2 |  |
| Post (M ± SD) | 83.2 ± 23.0 | 84.5 ± 17.2 | 82.2 ± 18.6 |  |

State Anxiety (STAI-S) change scores did not differ significantly between groups (F(2,57) = 0.60, p = 0.553, η² = 0.021), indicating no differential effect of the interventions on state anxiety as measured by the STAI-S. (sup fig.1).

Using the established Minimal Clinically Important Difference (MCID) of 10 points for VAS scales(39), responder analysis revealed clinically meaningful differences in treatment response. For anxiety reduction, VR+Meditation (68.2%) and VR Only (70.0%) (fig.2.e) showed similar effectiveness compared with control (27.8% Fisher’s exact p = .025 and .022, respectively), with NNT values of 2.5 and 2.4, respectively. For fear reduction, VR+Meditation demonstrated higher effectiveness (72.7% responders, NNT = 2.2, Fisher exact p = .010) compared to control (27.8%), while VR Only (45.0%, NNT =5.8) did not significantly differ from control (fig.2.f). For VAS Calm, although the omnibus ANOVA did not reach significance responder analysis only trended towards meaningful improvement in both VR+Meditation (50.0%, p = .046) and borderline significance in VR Only (55.0%, p = .020) versus control (16.7%), with NNTs of 3.0 and 2.6, respectively (fig.2 e-f) (table. 6). Optimistic response rates did not differ between groups. No significant differences were observed between VR+Meditation and VR Only on any measure (all p > .10). In summary, this responder analysis corroborates the group-level findings, confirming that both VR modalities deliver clinically meaningful anxiety relief and the addition of meditation shows a significant reduction in preoperative fear.

**Table.6.**

| **Measure** | **VR+Med**<br>**Responders** | **VR Only**<br>**Responders** | **Control**<br>**Responders** | **NNT:**<br>**VR+Med vs**<br>**Control** | **NNT:VR**<br>**Only vs**<br>**Control** |
| --- | --- | --- | --- | --- | --- |
| VAS<br>Anxious | 15/22 (68.2%) | 14/20 (70.0%) | 5/18 (27.8%) | 2.5 | 2.4 |
| VAS Afraid | 16/22 (72.7%) | 9/20 (45.0%) | 5/18 (27.8%) | 2.2 | 5.8 |
| VAS Calm | 11/22 (50.0%) | 11/20 (55.0%) | 3/18 (16.7%) | 3.0 | 2.6 |

To examine physiological changes we quantified heart rate and blood pressure difference scores. No significant between-group differences were detected for either systolic(F(2,56) = 0.93, p = 0.399; Kruskal-Wallis H = 0.81, p = 0.667) or diastolic blood pressure (*F*(2,55) = 0.54, p = 0.588; Kruskal-Wallis H = 0.78, p = 0.676). (sup fig 2.a-b).

### Secondary Outcomes – Physiological Arousal

Heart-rate change scores demonstrated significant between-group differences (F(2,57) = 3.86, p = .027, η² = .119; Kruskal–Wallis H = 17.05, p < .001). Given pronounced non-normality in the HR-change distributions, including one extreme value in the VR Only group (HR change = +53 bpm; pre = 66, post = 119 bpm), median-based descriptives and non-parametric tests are reported alongside means. Following the Kruskal–Wallis test, pairwise Mann–Whitney U tests with Bonferroni correction across the three comparisons showed significant differences between VR+Meditation and Care as Usual (U = 58.0, adjusted p < .001) and between VR Only and Care as Usual (U = 64.5, adjusted p = .002). The two VR conditions were not statistically distinguishable. VR+Meditation showed a consistent heart-rate reduction (median = −2.0 bpm, IQR [−8.8, −0.2]; mean = −3.5 bpm, SD = 7.0), confirmed by both a paired t test (t(21) = −2.38, p = .027) and Wilcoxon test (W = 55.5, p = .037). The control group showed a consistent increase (median = +4.5 bpm, IQR [2.0, 7.0]; mean = +5.0 bpm, SD = 4.1; t(17) = 5.14, p < .001; Wilcoxon W = 5.0, p < .001). These findings indicate that the standard preoperative waiting period was associated with increasing heart rate that was shown by the control group, this increase in physiological arousal was mitigated by the VR interventions.

The single extreme outlier in the VR-only group heart rate rose markedly (+53 bpm; pre = 66, post = 119 bpm). A sensitivity analysis excluding this participant yielded a mean reduction of −3.2 bpm (SD = 7.7), closely paralleling the VR+Meditation group, though remaining non-significant (t(18) = −1.79, p = 0.091). While a transcription error cannot be definitively excluded, concurrently recorded diastolic blood pressure for this participant rose by 17 mmHg (74 to 91 mmHg), suggesting a genuine physiological event; therefore, the value was retained in the primary analysis. Interestingly, this participant rated the VR experience as highly relaxing (85/100) and reported minimal cybersickness (nausea, dizziness, headache, and blurred vision), indicating no VR related adverse events.

Analysis of change scores showed that reductions in VAS Afraid were significantly correlated with heart rate reductions (r = 0.347, 95% CI [0.102, 0.552], p = 0.007) (fig.2.i), whereas VAS Anxious reductions showed no such relationship (r = 0.139, 95% CI [-0.119, 0.380], p = 0.288) (fig.2.j). This pattern did not differ significantly between treatment groups (Fisher’s Z comparisons: all p ≥ 0.27). This correlational data suggest that autonomic arousal is linked specifically with the alleviation of surgery-related fear, rather than anxiety in this setting.

To examine whether prior experience predicted treatment response, Pearson correlations were computed within relevant subgroups. Prior VR experience (M = 10.4, SD = 16.8) was examined within the two VR groups combined (n = 42) and prior meditation experience (M = 12.8, SD = 19.8) within the VR+Meditation group (n = 22). Neither was significantly correlated with any outcome change score (prior VR: all |r| ≤ 0.10, all p > 0.50; prior meditation: all |r| < 0.18, all p ≥ 0.44; S3 Fig a–d). Most patients had little or no prior experience (71% of VR-arm and 59% of VR+Meditation patients rated it below 10/100; S3 Fig e), which limits the power of these correlations. In an exploratory analysis of VR- and meditation-naïve patients, reductions relative to care as usual were similar in size to the full sample: VR-naïve patients in both VR groups (n = 30; anxiety d = −0.91, p < .001; fear d = −0.85, p = .003) and meditation-naïve patients in the VR+Meditation group (n = 13; anxiety d = −0.90, p = .039; fear d = −0.97, p = .028; S3 Fig f–g; Welch t-tests, unadjusted). This supports the use of both VR interventions for patients with little or no prior VR or meditation experience.

We conducted exploratory retrospective semi-structured interviews with patients during recovery to further validate the lower dimensional reports provided by visual analogue scales. A total of 135 state descriptors from 31 participants (fig 3.a-c) were analyzed in a mixed-effects model with condition as a fixed effect and participant as a random intercept. Relative to the Control group (n = 7), the VR+Meditation group (n = 11) showed significantly higher distress-relaxed projection scores (b = 0.123, SE = 0.050, p = .036), indicating greater alignment with the relaxed pole. The VR Only group (n = 13) did not differ significantly from Control (b = 0.062, SE=0.049, p=.234). Planned CR2-robust pairwise contrasts showed that VR+Meditation differed from Control (p = .042, with a moderate standardized effect (d = 0.51). The VR+Meditation versus VR Only contrast was not significant (p = .563), nor was the VR Only versus Control contrast (p = .602), all p-values are Bonferroni adjusted (fig 3.d).

**Figure 3.**
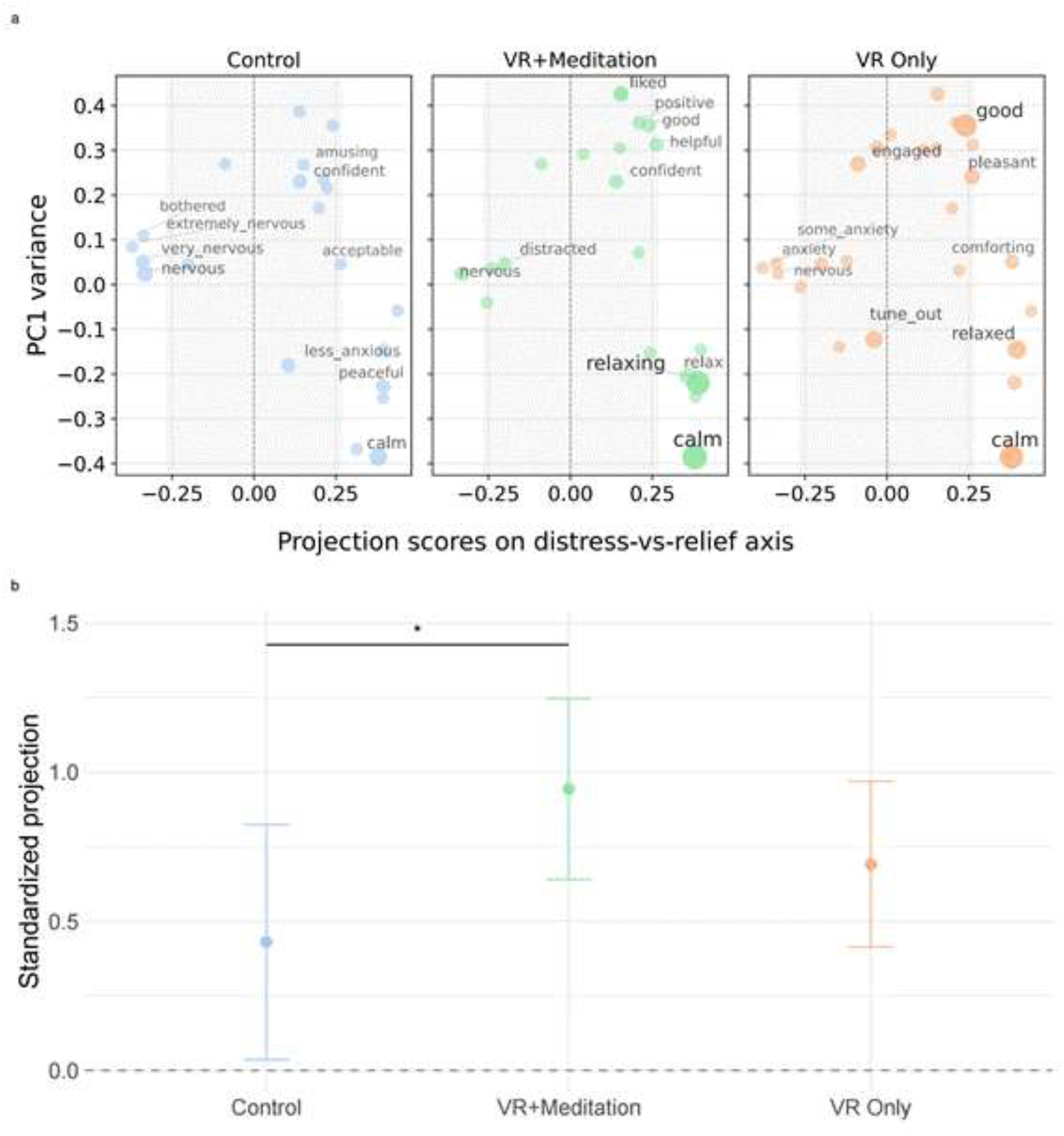
Qualitative semantic projection of post-operative patient interviews. **(a) Two-Dimensional Semantic Maps.** Scatter plots illustrating the distribution of interview state descriptors projected onto the shared distress-vs-relief axis (x-axis) and Principal Component 1 variance (y-axis) across the Control (blue), VR+Meditation (green), and VR Only (orange) cohorts. Labeled descriptors highlight distinct, group-specific state patterns (e.g., ‘extremely nervous’ clustering in Control vs. ‘relaxing’ and ‘calm’ clustering in the VR arms). The shaded vertical band marks the neutral region defined the median absolute deviation from zero (MAD0). Words falling to the left of this shaded region are distress-extreme, whereas words falling to the right are relief-extreme. In the primary analysis, distress-extreme words were most frequent in the Control condition (23.3%), compared with VR Only (5.3%) and VR+Meditation (4.2%), consistent with a significant overall Fisher test for the distress tail (p = 0.0157). **(b) Mean Standardized Projections**. Group-level comparison of the standardized mean projection scores derived from the semantic space. Dots represent group model-estimated means; error bars indicate 95% confidence intervals. The VR+Meditation cohort demonstrated a significantly higher alignment with relief sentiment compared to the Control group (*p<0.05). No significant differences were observed between the VR Only and Control groups, or directly between the two active VR arms.

As exploratory robustness checks, the word-level semantic projection pipeline was repeated using two alternative embedding spaces: *jinaai/jina-embeddings-v5-text-small*, representing a high-performing modern embedding model with a different architecture, and *glove-wiki-gigaword-300*, the original embedding family used by Grand et al. (2022) (38) for semantic projection. Across all three models, the primary VR+Meditation versus Control contrast remained significant after Bonferroni correction (Qwen: p = .042; Jina: p = .010; GloVe300: p = .005). Thus, the principal finding that the VR+Meditation condition was more strongly aligned with the relief pole than Control was robust across three distinct embedding spaces.

In an exploratory follow-up, semantically extreme words were identified using a zero-centered median absolute deviation (MAD0) threshold. Under this criterion, the Control condition showed the highest proportion of distress-extreme words (23.3%), whereas VR+Meditation showed the highest proportion of relief-extreme words (52.1%), with VR Only generally intermediate between the two. Consistent with this descriptive pattern, the overall Fisher test on extreme word counts was significant for distress-extreme word frequencies (p = .016) but not for relief-extreme word frequencies (p = .152).

Diagnostic checks indicated no singular fit and acceptable convergence, although heteroscedasticity and residual non-normality were detected; accordingly, inference was based on CR2 cluster-robust variance estimation rather than conventional model-based standard errors. No influential outliers were identified under the prespecified threshold, and explained variance was modest (marginal R^2 = .069, conditional R^2 = .104).

This exploratory analysis of patients’ retrospective account of their pre-surgical experience provides independent convergent evidence to the subjective reduction in fear and anxiety. Specifically that while VR alone provides moderate benefits across fear and anxiety, Meditation + VR significantly shifts the patients pre-surgical experience away from distress and towards one of relief relative to controls.

## Discussion

This randomized trial demonstrates that these brief, first time, immersive VR interventions are feasible, well tolerated, and associated with meaningful reductions in preoperative distress in patients awaiting breast cancer surgery. While both VR interventions reduced anxiety compared with standard care, the combination of nature VR and guided mindfulness was associated with a significant reduction in surgery-specific fear.

Although anxiety and fear are frequently considered together in perioperative research, they represent related but distinct psychological constructs. Generalized anxiety reflects a broader state of uncertainty and apprehension, whereas surgery-specific fear is directed toward the impending procedure itself. Our findings suggest that different immersive therapeutic content may target distinct dimensions of preoperative distress. Nature experiences may be sufficient to reduce generalized anxiety by immersing patients in a soothing environment distinct from the clinical context. The addition of guided mindfulness may help patients engage differently with surgery-specific fear, perhaps through regulation of attention.

Consistent with a substantial body of literature demonstrating the anxiolytic effects of both VR distraction and mindfulness, both active intervention groups significantly outperformed the CAU control group in reducing pre-operative anxiety as measured by Visual Analog Scales (VAS). Specifically, the VR Only group showed the largest effect size for anxiety reduction (Cohen’s d = −1.07), while the VR+Meditation group showed a large and significant reduction in both anxiety and, uniquely, in fear reduction (Cohen’s d = −0.99 relative to CAU). Recent meta-analyses of interventions in adult preoperative settings typically report small to moderate pooled effect sizes for reduction in pre-surgical distress with VR distraction (17,40); and in traditional, unassisted mindfulness interventions (8). The large effect sizes here are encouraging even given the small sample size. Responder analysis supports these benefits as being clinically meaningful. Both VR interventions provided comparable anxiety reduction (NNT = 2.4–2.5) and the addition of mindfulness provided a clinical advantage for mitigating surgery-related fear (NNT = 2.2 vs. 5.8 for VR Only). This distinction between the effect on VAS *anxiety* and *fear* suggests that different immersive interventions target different aspects of pre-surgical distress, in this case the added mindfulness component reducing fear associated with the impending surgical procedure.

Visual analogue scales are useful for rapid assessment of patients’ state, however they inevitably flatten patient experience into single data points. We conducted short interviews with the patients during recovery to gain a more nuanced understanding of the patient’s experiences in their own words. The analysis of these interviews support the findings from the VAS scales. The VR+meditation group uniquely shifted significantly toward the relief pole of the semantic axis compared to CAU. The VR+Meditation group utilized highly positive, regulation-focused language to describe the preoperative waiting time (e.g., ―relaxing‖, ―calm‖, ―positive‖, ―helpful‖). In contrast, the VR Only vocabulary was more broadly distributed, capturing the benefits of distraction (e.g., ―engaged‖, ―comforting‖, ―pleasant‖) but still including expressions of residual unease (e.g., ―some anxiety‖, ―nervous‖). Meanwhile, the control group used more distress-oriented descriptors (e.g., ―extremely nervous‖, ―very nervous‖, ―bothered‖). Importantly, this pattern emerged independently across patient-reported scales and computational analysis of patients’ own postoperative narratives, providing convergent evidence that the intervention influenced how participants experienced and subsequently described the preoperative period. Although exploratory, these findings suggest that computational analysis of patients’ spoken accounts may provide a valuable complementary approach for capturing psychological responses that are not fully reflected by conventional questionnaire-based measures.

The subjective and linguistic indicators of change in distress were supported by physiological changes. During the waiting period, heart rate increased in the CAU group while it decreased in both VR interventions indicating that VR was effective in managing sympathetic arousal. This aligns with prior work showing VR can reduce biomarkers of anxiety during acute medical procedures (41). Interestingly an exploratory correlation analysis indicated a significant relationship between change in heart rate and reported fear but not anxiety. These findings are consistent with the possibility that reductions in surgery-related fear and autonomic arousal are linked, although the present study cannot determine the direction or mechanism of this relationship. While environmental distraction alone (VR Only) can lower heart rate, the combined intervention may coordinate a coupling between physiological relaxation and reduction in pre-surgical fear.

Treatment effects were present in patients with little or no prior VR or meditation experience. The most effective mindfulness interventions tend to be those with prior patient training or with an in person practitioner as guide for first-time interventions. However despite variability in prior meditation experience among our patient cohort, this history did not predict treatment outcomes or moderate group differences. This was the same with experience with having no impact on outcomes, suggesting that the novelty of an immersive experience was not responsible for its therapeutic effect here. Together these findings support the viability of using VR both as distraction but particularly when combined with guided mindfulness exercises as an effective first-time intervention. This is particularly encouraging as VR devices are inexpensive, highly accessible and can be quickly deployed in the busy and fast paced perioperative context and with little training for staff.

The generalizability of these findings is limited by the small pilot sample. This may have undermined the ability to detect differences between the two VR conditions or led to inflated effect sizes that require a larger trial for confirmation. While we can compare VR+meditation to CAU and VR only, we did not have a fourth arm where mindfulness instruction was provided without VR, i.e. an instructional audio track alone, which means we cannot claim that virtual reality improves the efficacy of mindfulness instruction itself. Physiological monitoring was restricted to discrete pre- and post-intervention measurements preventing assessment of real-time autonomic trajectories or heart rate variability. Finally, this study did not track downstream post-operative outcomes, such as objective pain scores, analgesic consumption, or length of hospital stay. Larger, multi-arm randomized controlled trials utilizing continuous physiological tracking and longitudinal post-operative follow-up are required to confirm and extend these preliminary results.

This study is the first to compare the preliminary clinical efficacy of standard nature VR distraction with brief immersive VR mindfulness instruction during the adult pre-operative waiting period. It shows that both the standalone VR distraction and VR-guided meditation are feasible, well-tolerated, and effective first-time interventions for managing preoperative distress and associated physiological arousal in breast cancer patients. While both modalities successfully mitigate generalized state anxiety and lower heart rate, the addition of an active mindfulness component provides a therapeutic advantage by specifically reducing surgery-specific fear. This matched a psychological shift toward calmness and away from distress in the VR+meditation group that was detected in patients’ own recollective vocabulary during recovery. Beyond demonstrating feasibility this pilot study provides preliminary evidence that different digital interventions address distinct components of the patient experience. This suggests patient outcomes may be improved by targeting specific therapeutic content delivered in VR to the psychological needs of the patient.

## Data Availability

All relevant data are within the manuscript and its Supporting Information files. Patient's clinical data cannot be shared publicly because they are protected by law. Data are available from the Champalimaud Foundation clinical file system for researchers who meet the criteria for access to confidential data.

https://github.com/arsandru/preop-vr-NLP

https://github.com/SMckenzieRennie/preop-vr-analysis

## Author contributions

David Pinto: **Conceptualization, Methodology, Data Curation, Investigation, Project Administration, Resources, Supervision, Validation, Writing – Original Draft Preparation, Writing – Review & Editing**

Rogélio Andrès-Luna: **Conceptualization, Methodology, Data Curation, Investigation, Project Administration, Resources, Supervision, Validation, Writing – Review & Editing**

Adrian Razvan Sandru: **Conceptualization, Methodology, Resources, Data Curation, Formal Analysis, Software, Writing – Review & Editing**[5]

Lydia Fettweis-Netto: **Data curation, Writing - Review**[S6]

Carlos Mavioso: **Investigation**

Pedro Gouveia: **Investigation**

João Correia-Anacleto: **Investigation**

Luzia Travado: **Conceptualization**, **Methodology**

Patrícia Vaz: **Conceptualization**, **Methodology**

Alexandra Cruz: **Conceptualization**, **Methodology**

Zachary F Mainen: **Writing – Review & Editing**

Maria João Cardoso: **Investigation**, **Writing – Review & Editing**

Scott Rennie: **Conceptualization, Methodology, Data Curation, Formal Analysis, Investigation, Project Administration, Software, Supervision, Validation, Visualization, Writing – Original Draft Preparation, Review & Editing**[7]

## Supplementary figures

**S1 Fig.**
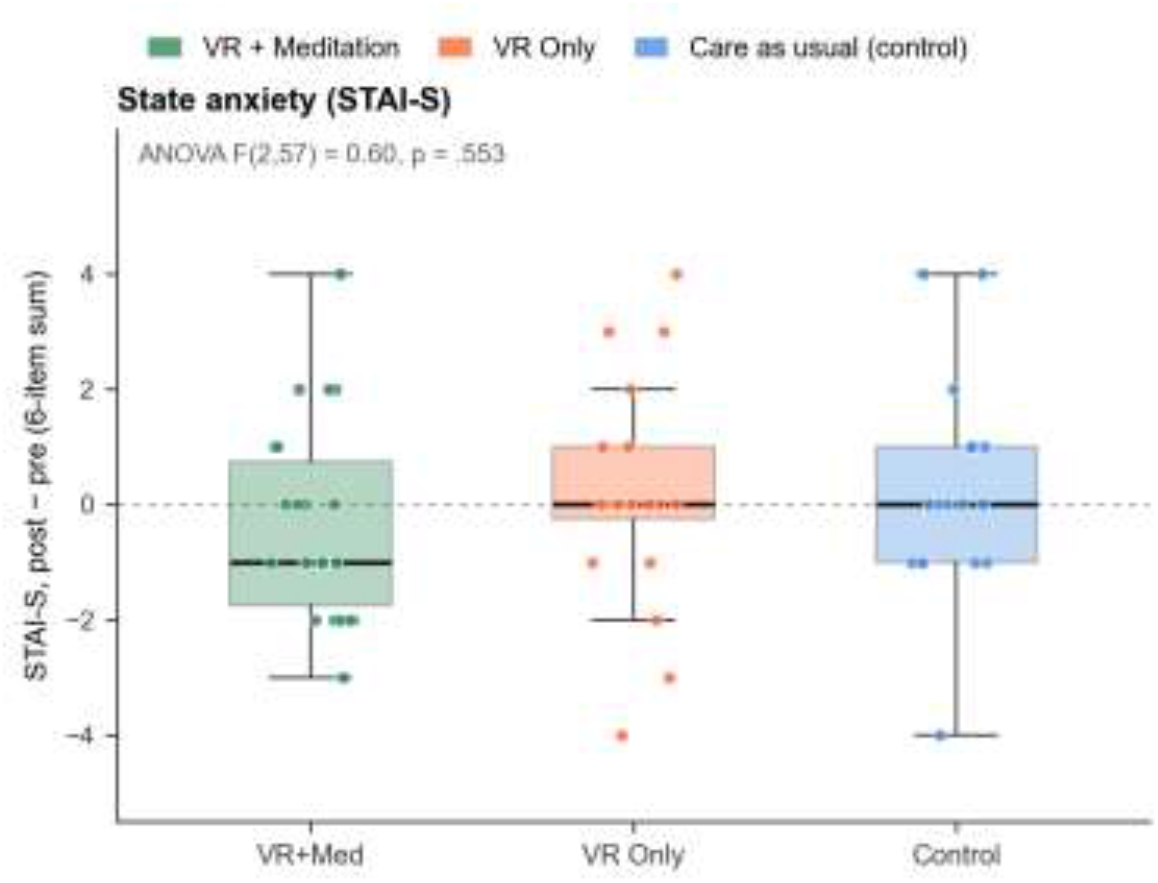
State anxiety (STAI-S) change scores. Box plots illustrating the change in STAI-S scores from baseline to post-intervention across the VR+Meditation, VR Only, and Control cohorts. Individual participant data points are overlaid. The horizontal dashed line at zero indicates no change from baseline; negative values represent a reduction in anxiety. A one-way ANOVA revealed no statistically significant differences in STAI-S change scores between the three groups.

**S2 Fig.**
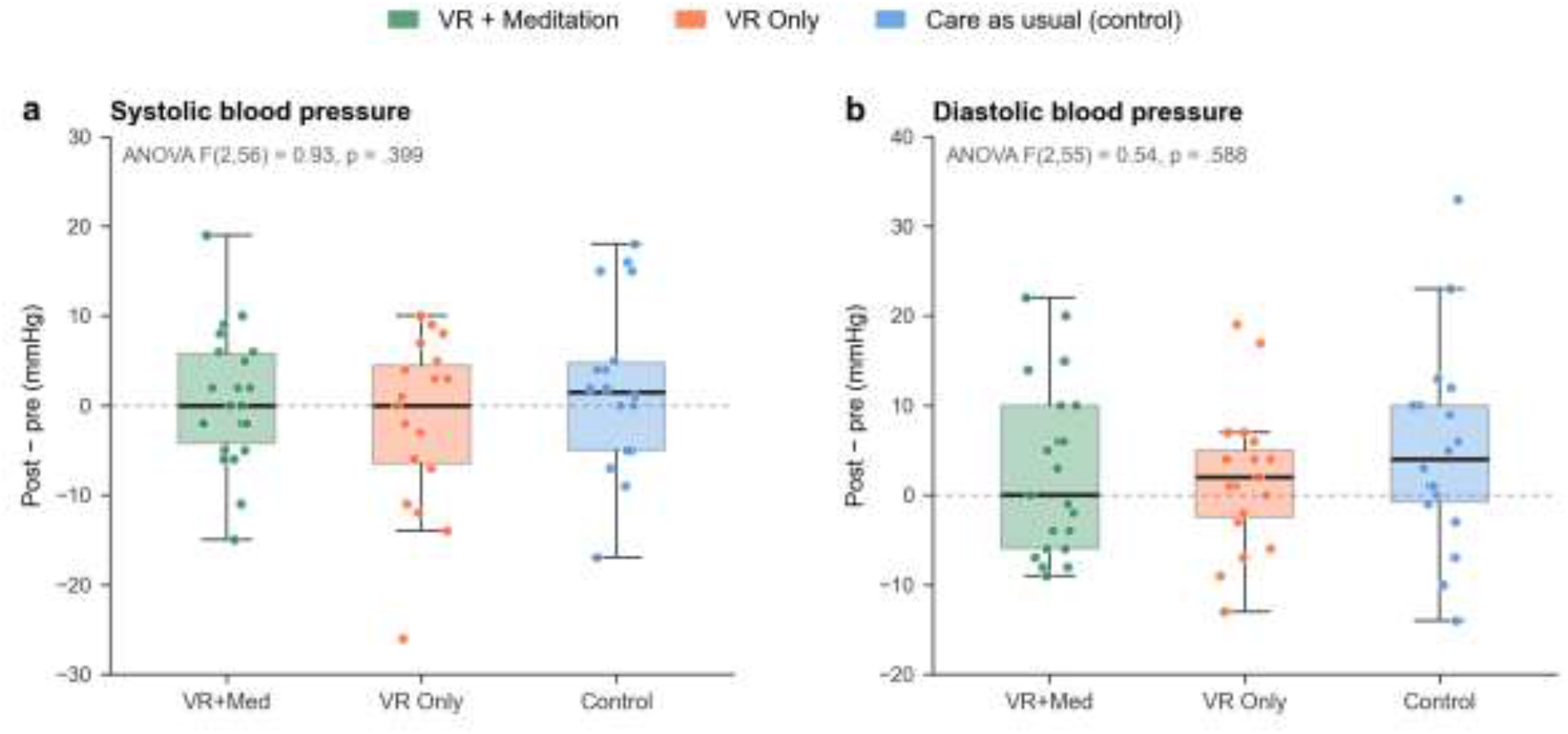
Blood pressure change scores. Box plots illustrating the change in systolic (left) and diastolic (right) blood pressure (mmHg) from baseline to post-intervention across the VR+Meditation, VR Only, and Control cohorts. Individual participant data points are overlaid. The horizontal dashed line at zero represents no change from baseline; negative values indicate a reduction in blood pressure.

**S3 Fig.**
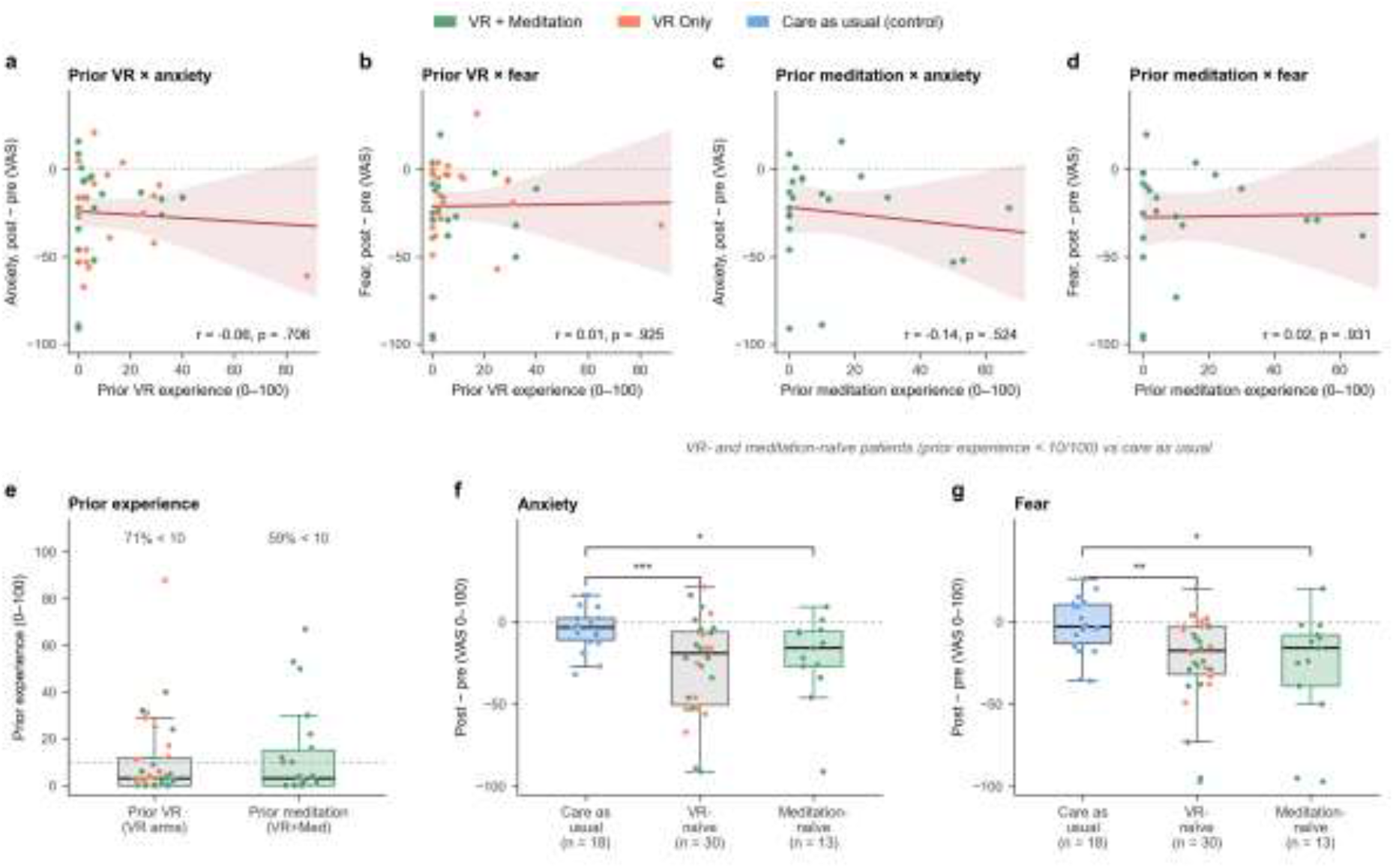
Prior experience and treatment response. (a–d) Self-reported prior experience (0–100) against pre-to-post change in anxiety and fear: prior VR experience in both VR arms (a, b; n = 42) and prior meditation experience in the VR+Meditation arm (c, d; n = 22). Lines show least-squares fits with 95% confidence bands; no correlation was significant. (e) Distribution of prior experience; the dashed line marks 10/100. (f, g) Anxiety (f) and fear (g) change compared with care as usual in VR-naïve patients (rating < 10, both VR arms; n = 30) and meditation-naïve patients (rating < 10, VR+Meditation arm; n = 13). Points are coloured by arm. *p < .05, **p < .01, ***p < .001, Welch t-test versus care as usual, unadjusted (exploratory).

